# Determinants of non-utilization of insecticide-treated nets among children under five in Rwanda: analyses of the 2024 Rwanda malaria indicator survey

**DOI:** 10.64898/2026.06.16.26355760

**Authors:** Kofi Nsaakoh Eduah, Abena Onormaah Eduah

## Abstract

**Background:** Insecticide-treated nets (ITNs) are effective for preventing malaria among children under five years, who bear a disproportionate burden of malaria. This study assessed the prevalence and determinants of ITN non-utilization among children under five in Rwanda using data from the 2024 Rwanda Malaria Indicator Survey (RMIS).

**Methodology:** This cross-sectional study utilized nationally representative data from the 2024 RMIS. Analyses were restricted to children under five residing in households that owned at least one ITN. The outcome was non-utilization of ITN, defined as not sleeping under an ITN the night preceding the survey. Survey-weighted descriptive statistics were used to estimate the prevalence of ITN non-utilization. Factors associated with non-utilization were identified using a survey-weighted Poisson regression model. Adjusted prevalence ratios (aPRs), 95% confidence intervals and p-values were reported.

**Results:** A total of 1,979 children were included in the study. The weighted prevalence of ITN non-utilization among children under five years was 20.11% (95% CI: 17.81 – 22.63). After adjusting for other factors, children aged 2 – 3 years were associated with an 83% higher prevalence of ITN non-utilization compared with those aged ≤1 year (aPR = 1.83, 95% CI: 1.423 – 2.352, p < 0.001). Compared with households that owned only one ITN, children in households with three or more ITNs were associated with a 76% lower prevalence of ITN non-utilization (aPR = 0.24, 95% CI: 0.171 – 0.332, p < 0.001). Children living in households with 5 – 7 members were associated with an 87% higher prevalence of ITN non-utilization compared with those in households with 1 – 4 members (aPR = 1.87, 95% CI: 1.476 – 2.358, p < 0.001).

**Conclusion:** The findings suggest that ITN utilization among children is influenced not only by household access to nets but also by household composition and dynamics that shape the allocation and use of available preventive resources.

## Introduction

Malaria remains one of the most important public health challenges globally, despite substantial progress in prevention and control over the last two decades. According to the World Health Organization (WHO), an estimated 263 million malaria cases and nearly 600,000 malaria-related deaths occurred worldwide in 2023, with the WHO African Region accounting for about 95% of deaths [1]. Children under five years of age continue to bear a disproportionate share of the malaria burden because of their relatively immature immunity and increased vulnerability to severe disease and death [2,3].

Among the recommended malaria prevention interventions, insecticide-treated nets (ITNs), particularly long-lasting insecticidal nets (LLINs), remain one of the most cost-effective and impactful vector-control strategies [4]. Evidence from systematic reviews and large-scale implementation programs has consistently demonstrated that ITNs substantially reduce human-vector contact, malaria incidence, severe malaria, and all-cause child mortality in endemic settings [5,6]. Consequently, universal access to and utilization of ITNs have remained central pillars of global and national malaria control strategies.

In sub-Saharan Africa, considerable investments have been made to increase household ownership and access to ITNs through mass distribution campaigns, antenatal care services, and routine child health programs [7,8]. Nevertheless, growing evidence suggests that ownership does not necessarily translate into utilization [9,10]. A systematic review of studies conducted across sub-Saharan Africa found substantial gaps between ITN ownership and actual use among children under five years, with utilization influenced by a complex interplay of individual, household, socioeconomic, environmental, and health-system factors [9,11]. Common determinants include caregiver education, household wealth, family size, access to malaria-related information, perceived malaria risk, sleeping arrangements, and the adequacy of ITN supply within households [12,13]. Barriers such as hot weather, perceived absence of mosquitoes, damaged nets, inadequate sleeping space, and low risk perception have also been reported to reduce utilization [14,15].

Rwanda has long been recognized as one of the countries that achieved notable gains in malaria control through the scale-up of integrated interventions, including widespread ITN distribution, indoor residual spraying, improved case management, and strengthened surveillance systems [16,17]. Between 2007 and 2010, Rwanda implemented large-scale malaria control efforts that contributed to significant reductions in malaria prevalence and under-five mortality [18]. In 2010 alone, approximately 4.1 million ITNs were distributed through a universal coverage campaign aimed at ensuring one net for every two persons and adequate coverage of household sleeping spaces [19].

Despite these achievements, malaria remains a persistent public health concern in Rwanda. Evidence indicates that after several years of decline, malaria cases have periodically resurged in different parts of the country, highlighting ongoing transmission risks and the need to sustain preventive interventions [20]. Spatial analyses of malaria among Rwandan children have demonstrated considerable geographical heterogeneity in malaria prevalence, with certain districts experiencing substantially higher risks than others due to environmental, climatic, and socioeconomic factors [21,22]. Such variations accentuate the importance of maintaining high coverage and consistent utilization of preventive measures, particularly among vulnerable populations such as children under five years of age.

The 2024 Rwanda Malaria Indicator Survey (RMIS) provides an important opportunity to examine current levels of ITN utilization among children under five and to identify factors associated with non-utilization in a nationally representative sample. Findings from such analyses can support the Malaria and Other Parasitic Diseases Division (MOPDD) which operates under the Rwanda Biomedical Centre (RBC), responsible for malaria prevention in Rwanda in developing targeted behavioural, educational, and community-based interventions aimed at maximizing the protective benefits of ITNs. Accordingly, this study assessed the determinants of non-utilization of ITNs among children under five years in Rwanda using data from the 2024 RMIS.

## Materials and methods

### Study design and data source

This study employed a cross-sectional analytical design using secondary data from the 2024 Rwanda Malaria Indicator Survey (RMIS). The RMIS is a nationally representative household survey conducted to monitor malaria-related indicators, including ownership and utilization of ITNs, malaria prevention practices, and other health-related outcomes among the Rwandan population. The survey was implemented using a stratified two-stage cluster sampling design, ensuring representation across all regions of Rwanda and by place of residence (urban and rural) [1].

The RMIS data were obtained from the Demographic and Health Surveys (DHS) Program database on 26/05/2026. The survey collected information from selected households through standardized questionnaires administered to eligible household members. The household recode and household member recode datasets were merged using unique household identification numbers to obtain both household and child-level information relevant to the study objectives. The present analyses focused on identifying factors associated with non-utilization of ITNs among children under five years of age in Rwanda. Given that the study utilized nationally representative survey data, findings can be generalized to children under five years living in households that owned at least one ITN in Rwanda during the survey period.

### Study sample and eligibility criteria

The study population comprised children aged below five years residing in surveyed households during the 2024 RMIS. Children were included in the analysis if they had slept in the household the night preceding the survey and were younger than five years of age. To specifically assess determinants of ITN non-utilization among children with access to mosquito nets, the analysis was restricted to children living in households that owned at least one insecticide-treated mosquito net. Household ITN ownership was determined from the household mosquito net inventory, and households without any ITNs were excluded from the analysis. There were no missing values on ITN utilization and all other factor variables. After applying all eligibility criteria and exclusions, a final sample of 1,979 children under five years of age was included in the analysis. These children constituted the analytical sample used to estimate the prevalence of ITN non-utilization and to examine its associated factors.

### Study variables

#### Outcome variable

The outcome variable was non-utilization of ITN among children under five years of age. This variable was derived from the survey question that assessed whether the child slept under an ITN on the night preceding the survey. Children who slept under an ITN the previous night were coded as 0 (used ITN), while those who did not sleep under an ITN were coded as 1 (did not use ITN). The outcome variable was analysed as a binary variable.

#### Explanatory variables

The explanatory variables were selected based on evidence from previous studies examining factors associated with ITN utilization among children [2–5] and the availability of relevant variables in the 2024 RMIS. The variables included age of the child (≤1 year, 2 – 3 years, and 4 years), sex of the child (male or female), place of residence (urban or rural), region (Kigali, South, West, North, and East), household wealth index, a standard DHS composite measure derived from household assets and housing characteristics (poorest, poorer, middle, richer, and richest), sex of the household head (male or female), number of ITNs in the household (1, 2, or ≥3 ITNs), household size (1 – 4, 5 – 7, or ≥8 members), household crowding (calculated by dividing the number of household members by the number of sleeping rooms; classified as not crowded [≤1 person per room], overcrowded [1.01 – 2 persons per room], and seriously overcrowded [>2 persons per room]), and relationship to the household head (child of head, grandchild, or other relationship).

### Statistical analyses

Data were analysed using Stata version 18 (StataCorp LLC, College Station, Texas, USA). The analysis accounted for the complex sampling design of the 2024 RMIS by applying sampling weights, primary sampling units, and stratification variables using the survey (svy:) commands. Sampling weights were generated by dividing the household sampling weight by 1,000,000. Descriptive statistics were used to summarize the characteristics of the study sample. Continuous variables were summarized using means and standard deviations, whereas categorical variables were described using frequencies and weighted percentages. The weighted proportion of ITN non-utilization among children under five years was estimated together with its 95% confidence interval (CI).

Bivariate analyses were performed using survey-weighted Poisson regression to examine the association between each explanatory variable and ITN non-utilization. Crude prevalence ratios (PRs), corresponding 95% confidence intervals, and p-values were reported. Variables assessed in the bivariate analysis included age and sex of the child, place of residence, region, household wealth index, sex of the household head, number of ITNs in the household, household size, household crowding, and relationship to the household head.

Subsequently, variables that demonstrated statistical significance at the bivariate analysis level (p < 0.05) were included in the survey-weighted multivariable Poisson regression model to identify factors independently associated with ITN non-utilization. Sex of the child was retained in the adjusted model irrespective of its bivariate significance because of its established epidemiological relevance. Adjusted prevalence ratios (aPRs) with their corresponding 95% confidence intervals were reported. Poisson regression was preferred because the outcome was binary and relatively common (20.11%), making prevalence ratios more appropriate and interpretable than odds ratios [6,7]. Multicollinearity among the explanatory variables was assessed using the variance inflation factor (VIF). The mean VIF was 1.87, indicating no evidence of problematic multicollinearity among the independent variables. Statistical significance was determined at a two-sided p-value of <0.05.

### Ethical considerations

This study was based on secondary analyses of anonymized data obtained from the 2024 RMIS through the Demographic and Health Surveys (DHS) Program. Permission to access and use the dataset was obtained from the DHS Program before the commencement of the analyses. The original survey protocols were reviewed and approved by the Malaria and Other Parasitic Diseases Division (MOPDD) of the Rwanda Biomedical Centre (RBC), which operates under the Ministry of Health, Rwanda and by the Institutional Review Board of ICF. Written informed consent was obtained from all eligible respondents before participation in the survey. The datasets made available by the DHS Program are fully de-identified and contain no information that could be used to identify individual participants. Therefore, no additional ethical approval or informed consent was required for this secondary data analyses.

## Results

### Background characteristics of study sample

A total of 1,979 children were included in the study, with a mean age of 1.99 years (SD = 1.45). Children aged ≤1 year constituted the largest proportion of the sample (n = 835; 42.24%), followed by those aged 2–3 years (n = 724; 36.83%) and 4 years (n = 420; 20.93%). The sex distribution was nearly equal, with females accounting for 50.49% (n = 986) and males 49.51% (n = 993). Most children resided in rural areas (n = 1,294; 68.02%), while 31.98% (n = 685) lived in urban settings. The highest proportion of participants were from the West region (n = 536; 27.35%), followed by the East (n = 299; 20.16%), South (n = 406; 19.77%), North (n = 397; 17.84%), and Kigali (n = 341; 14.88%).

Household wealth was almost evenly distributed across the wealth quintiles, with the largest proportion belonging to the poorer quintile (n = 425; 21.60%), followed by the richest (n = 449; 21.10%), richer (n = 403; 20.95%), middle (n = 388; 20.51%), and poorest (n = 314; 15.83%) quintiles.

Most households were headed by males (n = 1,422; 71.23%). The mean number of ITNs per household was 1.99 (SD = 0.99). Most of households owned one ITN (n = 710; 39.49%), while 35.82% (n = 709) owned two ITNs and 24.68% (n = 530) owned three or more ITNs. The mean household size was 4.79 members (SD = 1.73). Nearly half of the households had 1 – 4 members (n = 980; 49.77%), whereas 41.98% (n = 836) had 5–7 members and 8.25% (n = 163) had eight (8) or more members. Regarding household crowding, the mean crowding index was 2.16 persons per room (SD = 0.86). Nearly half of households were overcrowded (n = 988; 49.83%), while 40.85% (n = 801) were seriously overcrowded and only 9.32% (n = 190) were not crowded. Most children were biological children of the household head (n = 1,711; 87.20%), with smaller proportions being grandchildren (n = 220; 10.51%) or having other relationships to the household head (n = 48; 2.30%). Table 1 shows details of background characteristics of the study sample.

**Table 1.** Background characteristics of study sample.

| Variable | N = 1,979 | Weighted percentage (%) |
| --- | --- | --- |
| **Age of child (years)** |  |  |
| Mean (Standard deviation) | 1.99 (1.45) |  |
| ≤1 | 835 | 42.24 |
| 2 – 3 | 724 | 36.83 |
| 4 | 420 | 20.93 |
| **Sex of child** |  |  |
| Male | 993 | 49.51 |
| Female | 986 | 50.49 |
| **Place of residence** |  |  |
| Urban | 685 | 31.98 |
| Rural | 1,294 | 68.02 |
| **Region** |  |  |
| Kigali | 341 | 14.88 |
| South | 406 | 19.77 |
| West | 536 | 27.35 |
| North | 397 | 17.84 |
| East | 299 | 20.16 |
| **Household wealth index** |  |  |
| Poorest | 314 | 15.83 |
| Poorer | 425 | 21.60 |
| Middle | 388 | 20.51 |
| Richer | 403 | 20.95 |
| Richest | 449 | 21.10 |
| **Sex of household head** |  |  |
| Male | 1,422 | 71.23 |
| Female | 557 | 28.77 |
| **Number of ITNs in household** |  |  |
| Mean (Standard deviation) | 1.99 (0.99) |  |
| 1 | 710 | 39.50 |
| 2 | 709 | 35.82 |
| 3+ | 530 | 24.68 |
| **Household size (members)** |  |  |
| Mean (Standard deviation) | 4.79 (1.73) |  |
| 1 – 4 | 980 | 49.77 |
| 5 – 7 | 836 | 41.98 |
| 8+ | 163 | 8.25 |
| **Household crowding ¶** |  |  |
| Mean (Standard deviation) | 2.16 (0.86) |  |
| Not crowded | 190 | 9.32 |
| Overcrowded | 988 | 49.83 |
| Seriously overcrowded | 801 | 40.85 |
| **Relationship to household head** |  |  |
| Child of head | 1,711 | 87.20 |
| Grandchild | 220 | 10.51 |
| Other ¶¶ | 48 | 2.30 |

¶ Not crowded: ≤1 person/room; Overcrowded: 1.01 – 2 persons/room; Seriously overcrowded: >2
persons/room
¶¶Other includes: son/daughter-in-law; brother/sister; adopted/foster child; other relative & not related

### Weighted proportion of non-utilization of ITN among children under 5 years in Rwanda

The pie chart (Fig 1) illustrates ITN non-utilization among children under 5 years in Rwanda. Of the 1,979 children included, 386 children did not sleep under an ITN the previous night, corresponding to a weighted prevalence of 20.11% (95% CI: 17.81 – 22.63). Conversely, 1,593 children (79.89%; 95% CI: 77.37–82.19) were reported to have slept under an ITN the night before the survey.

**Fig 1.**
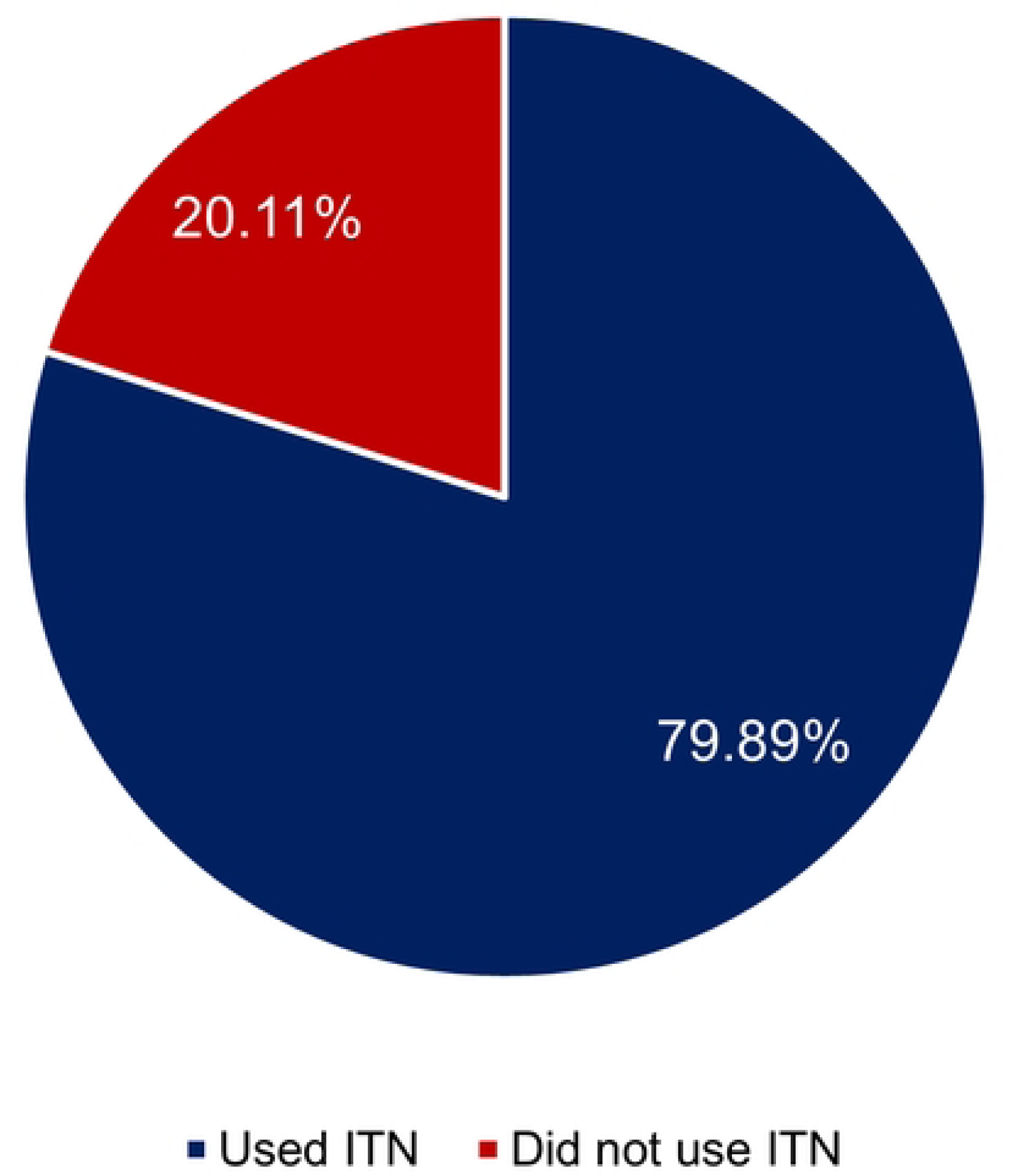
Weighted proportion of ITN non-utilization among children under 5 years in Rwanda.

### Bivariate Poisson regression analysis of factors associated with non-utilization of ITNs among children under 5 years in Rwanda

Table 2 presents the bivariate Poisson regression analysis of factors associated with non-utilization of ITNs among children under five years in Rwanda. Compared with children aged ≤1 year, children aged 2 – 3 years had a 77% higher prevalence of not using an ITN (PR = 1.77, 95% CI: 1.363 – 2.299, p < 0.001), while children aged 4 years had more than twice the prevalence of non-utilization of ITNs (PR = 2.31, 95% CI: 1.828 – 2.931, p < 0.001). Children living in rural areas had a 40% higher prevalence of not using an ITN compared with those residing in urban areas (PR = 1.40, 95% CI: 1.115 – 1.769, p = 0.004).

**Table 2.** Survey-weighted bivariate Poisson regression analyses of factors associated with non-utilization of ITNs among children under 5 years in Rwanda.

| Variable | Prevalence Ratio (PR) | 95% CI | p-value |
| --- | --- | --- | --- |
| **Age of child (years)** |  |  |  |
| ≤1 | Ref | – | – |
| 2 – 3 | 1.77 | 1.363 – 2.299 | <0.001* |
| 4 | 2.31 | 1.828 – 2.931 | <0.001* |
| **Sex of child** |  |  |  |
| Male | Ref | – | – |
| Female | 0.89 | 0.726 – 1.084 | 0.238 |
| **Place of residence** |  |  |  |
| Urban | Ref | – | – |
| Rural | 1.40 | 1.115 – 1.769 | 0.004* |
| **Region** |  |  |  |
| Kigali | Ref | – | – |
| South | 1.27 | 0.851 – 1.903 | 0.238 |
| West | 1.14 | 0.800 – 1.624 | 0.465 |
| North | 1.23 | 0.805 – 1.878 | 0.336 |
| East | 1.48 | 1.057 – 2.076 | 0.023* |
| **Household wealth index** |  |  |  |
| Poorest | Ref | – | – |
| Poorer | 0.86 | 0.596 – 1.229 | 0.396 |
| Middle | 0.81 | 0.572 – 1.160 | 0.253 |
| Richer | 0.75 | 0.526 – 1.075 | 0.117 |
| Richest | 0.72 | 0.520 – 0.997 | 0.048* |
| **Sex of household head** |  |  |  |
| Male | Ref | – | – |
| Female | 0.73 | 0.565 – 0.948 | 0.019* |
| **Number of ITNs in household** |  |  |  |
| 1 | Ref | – | – |
| 2 | 0.44 | 0.340 – 0.561 | <0.001* |
| 3+ | 0.43 | 0.318 – 0.590 | <0.001* |
| **Household size (members)** |  |  |  |
| 1 – 4 | Ref | – | – |
| 5 – 7 | 1.55 | 1.249 – 1.927 | <0.001* |
| 8+ | 1.81 | 1.267 – 2.593 | 0.001* |
| **Household crowding** |  |  |  |
| Not crowded | Ref | – | – |
| Overcrowded | 1.80 | 1.069 – 3.026 | 0.027* |
| Seriously overcrowded | 1.93 | 1.145 – 3.269 | 0.014* |
| **Relationship to household head** |  |  |  |
| Child of head | Ref | – | – |
| Grandchild | 1.28 | 0.960 – 1.707 | 0.091 |
| Other | 1.75 | 1.132 – 2.710 | 0.012* |

\*Statistically significant at p-value <0.05; Ref: Reference category

Regarding region, children residing in the Eastern region had a 48% higher prevalence of ITN non-utilization than those in Kigali (PR = 1.48, 95% CI: 1.057 – 2.076, p = 0.023). Children from the richest households had a 28% lower prevalence of ITN non-utilization compared with those from the poorest households (PR = 0.72, 95% CI: 0.520 – 0.997, p = 0.048). Children living in female-headed households were associated with a 27% lower prevalence relative to those in male-headed households (PR = 0.73, 95% CI: 0.565 – 0.948, p = 0.019).

Relative to households owning only one ITN, children in households with two ITNs had a 56% lower prevalence of non-utilization (PR = 0.44, 95% CI: 0.340 – 0.561, p < 0.001), while those in households with three or more ITNs had a 57% lower prevalence (PR = 0.43, 95% CI: 0.318 – 0.590, p < 0.001). Children living in households with 5 – 7 members had a 55% higher prevalence of non-utilization (PR = 1.55, 95% CI: 1.249 – 1.927, p < 0.001), whereas those in households with 8 or more members had an 81% higher prevalence (PR = 1.81, 95% CI: 1.267 – 2.593, p = 0.001), compared with children in households containing 1 – 4 members. Children in overcrowded households had an 80% higher prevalence of ITN non-utilization (PR = 1.80, 95% CI: 1.069 – 3.026, p = 0.027), while those in seriously overcrowded households had a 93% higher prevalence (PR = 1.93, 95% CI: 1.145 – 3.269, p = 0.014), relative to children in non-crowded households. With respect to relationship to the household head, children classified as other relatives had a 75% higher prevalence of ITN non-utilization than biological children of the household head (PR = 1.75, 95% CI: 1.132 – 2.710, p = 0.012). Sex of the child was not significantly associated with ITN non-utilization.

### Survey-weighted multivariable Poisson regression analyses of factors associated with non-utilization of ITNs among children under 5 years in Rwanda

Table 3 presents the multivariable Poisson regression analysis of factors independently associated with non-utilization of ITNs among children under five years in Rwanda. After adjusting for other factors in the model, children aged 2 – 3 years were associated with an 83% higher prevalence of ITN non-utilization compared with children aged ≤1 year (aPR = 1.83, 95% CI: 1.423–2.352, p < 0.001), while children aged 4 years had 2.55 times the prevalence of ITN non-utilization compared with children aged ≤1 year (aPR = 2.55, 95% CI: 2.011–3.231, p < 0.001). Children living in female-headed households had a 23% lower prevalence of ITN non-utilization compared with those living in male-headed households (aPR = 0.77, 95% CI: 0.595–0.990, p = 0.042).

**Table 3.** Survey-weighted multivariable Poisson regression analysis of factors associated with non-utilization of ITNs among children under 5 years in Rwanda.

| Variable | Adjusted-Prevalence Ratio (aPR) | 95% CI | p-value |
| --- | --- | --- | --- |
| **Age of child (years)** |  |  |  |
| ≤1 | Ref | – | – |
| 2 – 3 | 1.83 | 1.423 – 2.352 | <0.001* |
| 4 | 2.55 | 2.011 – 3.231 | <0.001* |
| **Sex of child** |  |  |  |
| Male | Ref | – | – |
| Female | 0.88 | 0.718 – 1.072 | 0.197 |
| **Place of residence** |  |  |  |
| Urban | Ref | – | – |
| Rural | 1.30 | 0.964 – 1.745 | 0.086 |
| **Region** |  |  |  |
| Kigali | Ref | – | – |
| South | 0.92 | 0.557 – 1.530 | 0.755 |
| West | 1.13 | 0.701 – 1.811 | 0.619 |
| North | 1.29 | 0.777 – 2.141 | 0.321 |
| East | 1.01 | 0.624 – 1.630 | 0.972 |
| **Household wealth index** |  |  |  |
| Poorest | Ref | – | – |
| Poorer | 0.83 | 0.591 – 1.157 | 0.265 |
| Middle | 0.84 | 0.603 – 1.184 | 0.325 |
| Richer | 0.88 | 0.614 – 1.272 | 0.504 |
| Richest | 1.07 | 0.758 – 1.510 | 0.697 |
| **Sex of household head** |  |  |  |
| Male | Ref | – | – |
| Female | 0.77 | 0.595 – 0.990 | 0.042* |
| **Number of ITNs in household** |  |  |  |
| 1 | Ref | – | – |
| 2 | 0.36 | 0.278 – 0.457 | <0.001* |
| 3+ | 0.24 | 0.171 – 0.332 | <0.001* |
| **Household size (members)** |  |  |  |
| 1 – 4 | Ref | – | – |
| 5 – 7 | 1.87 | 1.476 – 2.358 | <0.001* |
| 8+ | 2.65 | 1.921 – 3.652 | <0.001* |
| **Household crowding** |  |  |  |
| Not crowded | Ref | – | – |
| Overcrowded | 1.60 | 0.963 – 2.664 | 0.069 |
| Seriously overcrowded | 1.31 | 0.772 – 2.230 | 0.313 |
| **Relationship to household head** |  |  |  |
| Child of head | Ref | – | – |
| Grandchild | 1.38 | 1.035 – 1.832 | 0.028* |
| Other | 1.79 | 1.172 – 2.740 | 0.007* |

Relative to households owning only one ITN, children in households with two ITNs were associated with a 64% lower prevalence of ITN non-utilization (aPR = 0.36, 95% CI: 0.278–0.457, p < 0.001), while those in households with three or more ITNs were associated with a 76% lower prevalence (aPR = 0.24, 95% CI: 0.171–0.332, p < 0.001). Compared with children living in households containing 1 – 4 members, those living in households with 5 – 7 members were associated with an 87% higher prevalence of ITN non-utilization (aPR = 1.87, 95% CI: 1.476–2.358, p < 0.001), whereas children in households with 8 or more members had 2.65 times the prevalence of ITN non-utilization compared with those living in households containing 1 – 4 members (aPR = 2.65, 95% CI: 1.921 – 3.652, p < 0.001). With respect to relationship to the household head, grandchildren were associated with a 38% higher prevalence of ITN non-utilization compared with biological children of the household head (aPR = 1.38, 95% CI: 1.035 – 1.832, p = 0.028). Similarly, children classified as other relatives were associated with a 79% higher prevalence of ITN non-utilization (aPR = 1.79, 95% CI: 1.172 – 2.740, p = 0.007). Sex of the child, place of residence, region, household wealth index, and household crowding were not significantly associated with ITN non-utilization after adjustment for other variables in the model.

## Discussion

One in five children did not sleep under an ITN the night preceding the survey, indicating that despite substantial progress in ITN distribution and ownership in Rwanda [16], gaps in utilization remain among a notable proportion of vulnerable children. This can potentially explain the persistence of barriers beyond access to ITNs and emphasize the importance of understanding household and child-level factors influencing net use. One of the most important findings of this study was the strong association between child age and ITN non-utilization. Increasing child age was independently associated with a higher prevalence of ITN non-utilization. This finding may suggest that younger children may be prioritized for malaria prevention within households because caregivers perceive infants and younger children as being more vulnerable to malaria-related morbidity and mortality [29]. As children grow older, caregivers may become less vigilant in supervising sleeping arrangements and ensuring consistent ITN use [30]. Older children may also sleep independently or in separate sleeping spaces, reducing caregiver oversight and increasing the likelihood of inconsistent ITN utilization [12,31]. Similar age-related disparities in ITN utilization have been reported in several African settings, where younger children were more likely to be protected by ITNs than older children [32,33]. Given the substantial magnitude of the association observed in this study, malaria control programmes must emphasize that all children under five remain highly vulnerable to malaria and should consistently sleep under ITNs regardless of age.

Children residing in households with more ITNs were less likely to experience non-utilization, emphasizing the potential role of adequate household net availability in promoting consistent use. Greater ITN availability improves the likelihood that all household members, including young children, have access to a sleeping space covered by a net. This finding is consistent with evidence from Rwanda and other sub-Saharan African countries showing that sufficient household ITN supply is one of the strongest predictors of utilization among children and other vulnerable populations [19,34,35]. The findings therefore reinforce the importance of sustaining universal coverage campaigns and routine ITN distribution strategies that ensure households possess enough nets relative to household size rather than merely owning at least one net.

Children residing in households with 5 – 7 members and those living in households with 8 or more members were more likely to not use ITNs compared with children from smaller households. Notably, the magnitude of the association increased with household size, suggesting a dose-response relationship. Larger households often experience greater pressure on available resources, including sleeping spaces and ITNs [36]. Even in households that own multiple nets, the number of household members may exceed the available net coverage, resulting in unequal allocation and lower utilization among some children [10]. An important observation from the present study is that household crowding was no longer statistically significant after adjustment, whereas household size remained independently associated with non-utilization. This finding could imply that the number of household members may be a more important determinant of ITN utilization than room occupancy itself. Otherwise stated, competition for available ITNs appears to be driven primarily by household composition rather than physical crowding alone.

An independent association was observed between household headship and ITN utilization, with children living in female-headed households exhibiting a lower prevalence of ITN non-utilization than their counterparts in male-headed households. Although the effect size was modest, the finding may reflect differences in household dynamics related to childcare and health-seeking behaviours. Female household heads may be more closely involved in decisions regarding children’s daily care and preventive health practices, including ITN use [37]. Similar findings have been reported in some studies conducted in sub-Saharan Africa, where female household leadership was associated with improved uptake of child health interventions and preventive practices [38,39].

Grandchildren and children categorized as other relatives were more likely to not use ITNs than biological children of the household head. This finding points to a potentially overlooked dimension of vulnerability within households. In situations where resources such as sleeping spaces or ITNs are limited, biological children may receive priority over grandchildren, foster children, or other dependent relatives [40]. Differences in caregiver attention, supervision, and resource allocation may contribute to disparities in ITN use among children residing within the same household [41]. This suggests that interventions promoting equitable allocation of preventive resources within households should specifically consider the needs of non-biological children and other dependent relatives who may be at increased risk of exclusion from malaria prevention practices.

Several factors that were significantly associated with ITN non-utilization in the bivariate analysis, including place of residence, region, household wealth, and household crowding, lost statistical significance after adjustment for other covariates. This suggests that their apparent effects were likely explained by underlying household characteristics, particularly ITN availability and household size. The loss of significance for wealth further indicates that socioeconomic differences in ITN utilization may operate indirectly through factors such as access to ITNs and living conditions rather than economic status itself. Similarly, the attenuation of the association between household crowding and ITN non-utilization after adjustment suggests that the number of household members may be more important than room occupancy in determining net use. Furthermore, the absence of a significant association between child sex and ITN utilization indicates that ITN use did not differ between male and female children in this population.

### Implications for practice

The findings of this study have important programmatic implications. Efforts to increase ITN utilization among children under five in Rwanda should move beyond household ownership indicators and prioritize universal access within households. Distribution strategies should ensure adequate net-to-person ratios, particularly for larger households. Behaviour change communication interventions should emphasize the importance of consistent ITN use among all children under five, including older children who may be overlooked despite remaining highly susceptible to malaria. Additionally, malaria prevention initiatives should ensure that behaviour change messaging and household-level ITN utilization strategies emphasize equitable ITN use among all children within a household, including grandchildren and other non-biological children. Collectively, these measures could contribute to reducing residual gaps in ITN utilization and strengthen malaria prevention among one of the most vulnerable population groups in Rwanda.

### Strengths and limitations of study

The study benefited from a relatively large sample size, which provided adequate statistical power to detect factors significantly associated with ITN non-utilization. The use of survey-weighted analyses accounted for the complex sampling design of the RMIS, ensuring nationally representative estimates and reducing the likelihood of biased inferences. Additionally, the application of multivariable Poisson regression allowed for the estimation of adjusted prevalence ratios, which are more appropriate and interpretable than odds ratios for common outcomes in cross-sectional studies. However, the cross-sectional nature of the RMIS data precludes the establishment of causal relationships between the identified factors and ITN non-utilization. Also, ITN utilization was measured based on caregiver self-report of whether the child slept under an ITN the night preceding the survey. Such information may be subject to social desirability bias, potentially leading to overreporting of ITN use. Finally, because ITN utilization was assessed for only the night preceding the survey, the measure may not fully reflect habitual or long-term net-use behaviours among children.

## Conclusion

Despite substantial progress in insecticide-treated net distribution in Rwanda, a considerable proportion of children under five years remain unprotected due to non-utilization. This study found that ITN non-utilization was associated with child age, household ITN availability, household size, household headship, and the child’s relationship to the household head. Collectively, these findings suggest that utilization of ITNs among children is shaped not only by access to nets but also by household structure and dynamics that influence the allocation and use of available preventive resources. Understanding these factors is important for informing efforts aimed at improving equitable and consistent ITN utilization among children under five years, a population that remains disproportionately vulnerable to malaria.

## Data Availability

No data were generated in this study. The data underlying the findings are available from the Demographic and Health Surveys (DHS) Program upon reasonable request and with permission from the data custodian. The 2024 Rwanda Malaria Indicator Survey dataset can be accessed through the DHS Program website (https://dhsprogram.com/data/) following registration and approval of a data access request.

## Acknowledgements

The authors would like to acknowledge the Demographic and Health Surveys (DHS) Program for granting access to the 2024 Rwanda Malaria Indicator Survey dataset used in this study. The author is also grateful to the survey participants and the institutions involved in the design, implementation, and management of the survey.

